# Is glucose-6-phosphate dehydrogenase deficiency associated with COVID-19 infection, severity, and death? A cohort study from the Brazilian Amazon

**DOI:** 10.1101/2025.08.21.25334198

**Authors:** Adila L B Dias, Joabi Rocha Nascimento, José Diego Brito-Sousa, Marco Aurelio Sartim, Kamilla Freitas da Silva, Alexandre Vilhena Silva-Neto, Gabriel dos Santos Mouta, Patricia Carvalho da Silva Balieiro, Djane Clarys Baia-da-Silva, Fernando Almeida-Val, Gisely Cardoso de Melo, Vanderson de Souza Sampaio, Marcus Lacerda, Wuelton Monteiro

## Abstract

**Background:** Glucose-6-phosphate dehydrogenase deficiency (G6PDd) is a common genetic disorder that impairs the cellular antioxidant response and has been hypothesized as a potential risk factor for severe outcomes in viral infections, including COVID-19. However, clinical evidence remains limited, especially in regions with high G6PDd prevalence.

**Methods:** We conducted a retrospective cohort study using secondary data from four health information systems from health facilities in the Brazilian Amazon (E-SUS Notifica, SIVEP-Gripe, SIM, and a G6PD enzyme activity database). The study population consisted of 3,955 male participants, including 206 with confirmed G6PDd. We used logistic regression to assess associations between G6PDd and COVID-19 infection, hospitalization, and death. Cox proportional hazards models and Kaplan-Meier curves were applied to evaluate time-to-event outcomes.

**Results:** No statistically significant association was found between G6PDd and SARS-CoV-2 infection (OR = 1.15; 95% CI: 0.70–1.79; p = 0.6), hospitalization (OR = 1.13; 95% CI: 0.27–3.20; p = 0.8), or death (OR = 0.00; p > 0.9). Age was a significant risk factor for all outcomes, and individuals identified as Asian had a higher likelihood of infection (OR = 2.87; 95% CI: 1.57–5.29; p < 0.001).

**Conclusion:** In this cohort from the Brazilian Amazon, G6PD deficiency was not associated with an increased risk of COVID-19 infection or severe outcomes. These findings emphasize the importance of considering ethnic and genetic diversity in epidemiological analyses and health policy planning, particularly in regions with high G6PDd prevalence.

## INTRODUCTION

The COVID-19 pandemic, caused by SARS-CoV-2, has had a major global impact since its emergence in December 2019 in Wuhan, China [1–3]. The World Health Organization (WHO) declared a Public Health Emergency of International Concern on January 30, 2020, and officially characterized it as a pandemic on March 11 of the same year [4,5]. In Brazil, the pandemic was particularly severe, with approximately 39.3 million confirmed cases and more than 700,000 deaths [6]. The state of Amazonas was one of the most affected regions, reporting 65,173 confirmed cases and 14,556 deaths, and facing a dramatic collapse in healthcare infrastructure, especially in Manaus, due to shortages of oxygen and hospital beds [7].

COVID-19 manifests with a broad clinical spectrum, from asymptomatic infection to severe illness, including pneumonia, respiratory failure, and multiorgan dysfunction [8–10]. Approximately 32% of SARS-CoV-2 infections are asymptomatic, and 20% of people remain symptom-free throughout the infection [11,12]. Among symptomatic individuals, 80% presented mild or moderate illness, 15% developed severe disease, and 5% progressed to critical illness with severe complications [11,13]. Several demographic and clinical factors influence disease severity, notably advanced age and the presence of preexisting comorbidities. Among these factors, genetic variations may also play a role during COVID infection, and some polymorphisms have been found to protect or outcomes.

G6PD is a key enzyme in the pentose phosphate pathway and is used to protect cells from oxidative stress [14,15]. G6PD deficiency, affecting over 400 million people worldwide, is characterized by reduced enzymatic activity and increased vulnerability to oxidative damage and hemolysis. This condition is a common enzymopathy linked to the Xq28 locus of the X chromosome. Although it is classified as recessive, both homozygous and heterozygous individuals may exhibit symptoms [16,17]. In Brazil, the estimated prevalence is around 5%, primarily due to the African A−variant [18]. Preclinical and clinical studies have shown that G6PDd, whether congenital or acquired, may be associated with increased or decreased vulnerability to SARS-CoV-2 infection [19–21]. While studies have shown that G6PD deficiency can also exacerbate specific aspects of COVID-19, such as respiratory dysfunction, anemia, and hepatic impairment [22], others found no association between G6PDd and mortality or need for ventilatory support [23]. Therefore, the clinical relationship between G6PD deficiency and COVID-19 severity is controversial and requires further investigation.

Health databases such as E-SUS Notifica, SIVEP-Gripe, and the Mortality Information System (SIM) are valuable tools for epidemiological surveillance and research in Brazil. These systems allow for the collection and integration of large-scale clinical and demographic data, contributing to evidence-based decision-making during pandemics [24–27].

This study aimed to investigate the association between G6PD deficiency and the incidence, hospitalization, and mortality due to COVID-19 in a male cohort from the Brazilian Amazon. Given the regional prevalence of G6PDd and the socioeconomic and ethnic diversity of the population, understanding this relationship may inform clinical management and public health strategies.

## MATERIALS AND METHODS

### Study Design and Setting

This was a retrospective cohort study (January 1, 2020 – December 31, 2022), conducted using clinical, demographic, epidemiological, and outcome data from Brazilian national databases: **e-SUS Notifica**, which records confirmed COVID-19 cases; **SIVEP-Gripe**, which includes information on COVID-19-related hospitalizations; and **SIM** (Mortality Information System), which contains records of deaths related to COVID-19 **(Figure 1)**. A database containing G6PD enzyme activity data from patients was compiled from three separate studies that evaluated G6PD status in distinct populations. Identification data from participants were accessed to link all the datasets and were deleted as soon as the final dataset was built. All the datasets were accessed on July 1, 2023. For this study, only male individuals were included in the analyses. The methods used to identify G6PD deficiency are described in the original papers [28–32]. The study was approved by the Research Ethics Committee (CAAE: 45714621.2.0000.0005), and data were obtained from different sources, including public health information systems from the state of Amazonas, Brazil, heavily affected by COVID-19 [7].

**Figure 1.**
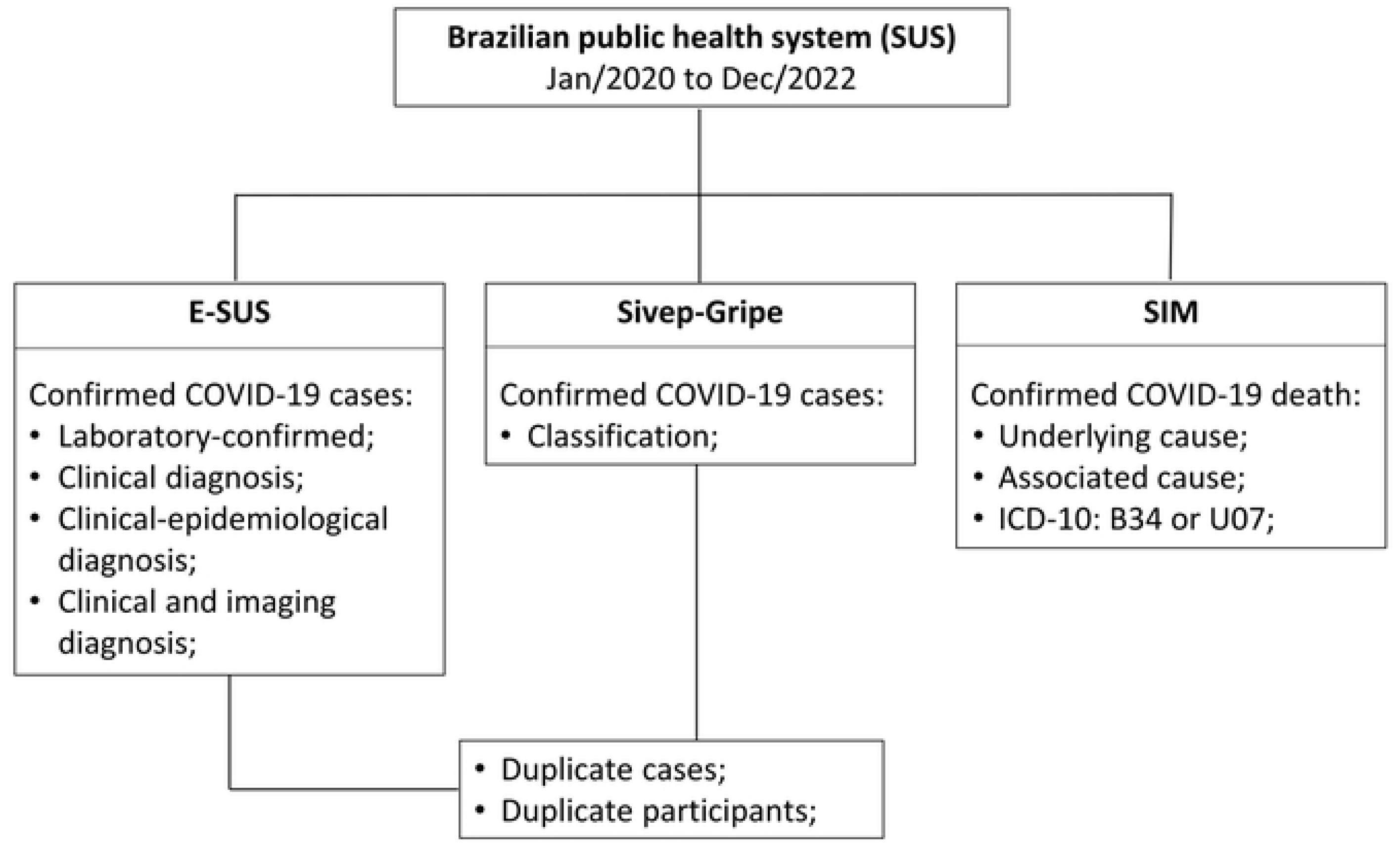
Flowchart describing data collection from the Brazilian Unified Health System (SUS) information systems.

### Data Processing: Deduplication and Record Linkage

The databases did not contain unique identifiers that would allow deterministic linkage. Therefore, they underwent a standardization process that included the removal of special characters, accents, and prepositions from participant names, as well as the formatting of birth dates. When birth dates were unavailable, recorded age was used for adjustment.

To remove redundant records, probabilistic deduplication was performed using the RecordLinkage package in R. The command RLBigDataDedup was used with the variables “Participant Name,” “Mother’s Name,” and “Date of Birth”; age was used as a substitute when the date of birth was missing. Next, probabilistic linkage between databases was performed to associate G6PD deficiency data with COVID-19, hospitalization, and death records. The linkage used the R programming language (RStudio 4.2.1) and the RecordLinkage library, employing the Levenshtein phonetic algorithm and blocking by sex. Identified pairs were manually reviewed by experts, who determined the match cutoff score.

### Data Analysis

The main explanatory variable was G6PD deficiency. In this study, participants with intermediate enzymatic activity were classified as having normal levels. The outcomes analyzed were confirmed COVID-19 infection, hospitalization, ICU admission, and death. Confirmed cases were identified in the SIVEP-Gripe and E-SUS Notifica databases. Hospitalizations were identified in SIVEP-Gripe, and deaths were included if COVID-19 was listed as the underlying or associated cause in the SIM.

Logistic regression was used to analyze associations between G6PD deficiency, ethnicity, age, and the outcomes (p < 0.05). Pearson’s chi-squared test was used for categorical variables, and the Wilcoxon test for continuous variables. Additionally, survival analysis was conducted to assess the association between G6PD deficiency and SARS-CoV-2 infection, time to hospitalization, ICU admission, and death, using the Cox proportional hazards model. Kaplan–Meier curves were used to estimate survival probabilities, and the log-rank test was used to compare groups.

All analyses were performed in R (version 4.4) and RStudio (2024.09.01), using the tidyverse, survival, and survminer packages. Graphs included 95% confidence intervals, and a 5% significance level was adopted.

## RESULTS

The study sample included 3,955 male participants, of whom 206 (5.2%) had G6PD deficiency (**Figure 2**). The demographic characteristics of participants are presented in **Table 1**. The mean age was very symmetrical among participant groups, with 35 years for both the total and Normal G6PD groups, and 34 years for the G6PD-deficient group. Regarding race, participants who self-reported admixed ethnicity consisted of the majority in all groups (over 72%); however, a noticeable distinction was observed in G6PD deficiency among Black (7.77%) and Asian (1.46%) participants.

**Figure 2.**
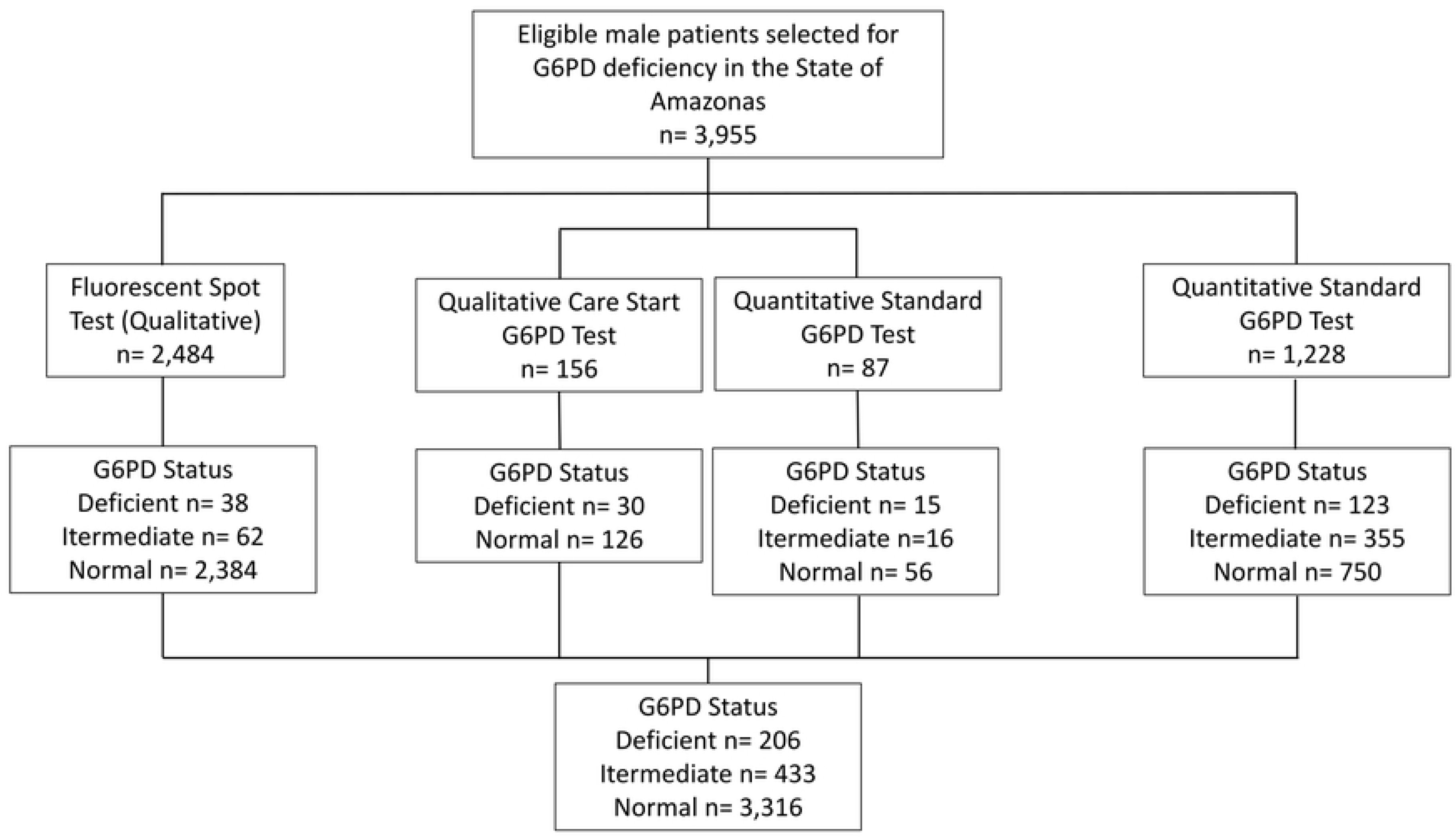
Flowchart describing the samples from the studies that compose the G6PD deficiency database.

**Table 1.** Demographic and clinical characteristics of study participants.

| Characteristics | Total<br>N = 3,955 | Normal<br>N = 3,749 | G6PD deficient<br>N = 206 |
| --- | --- | --- | --- |
| **Age, mean (SD)** | 35 (17) | 35 (17) | 34 (18) |
| **Ethnicity, n (%)** |  |  |  |
| White | 259 (6.55%) | 245 (6.54%) | 14 (6.80%) |
| Black | 612 (15.47%) | 596 (15.90%) | 16 (7.77%) |
| Asian | 125 (3.16%) | 122 (3.25%) | 3 (1.46%) |
| Admixed | 2,899 (73.30%) | 2,729 (72.79%) | 170 (82.52%) |
| Indigenous | 60 (1.52%) | 57 (1.52%) | 3 (1.46%) |

In the multivariate analysis, no statistically significant association was observed between G6PD deficiency and SARS-CoV-2 infection (OR = 1.15; 95% CI: 0.70-1.79; p = 0.6) (**Table 2**). Age was significantly associated with infection (OR = 1.02; p < 0.001), as was Asian (OR = 2.87; 95% CI: 1.57-5.29; p < 0.001).

**Table 2.** Association between G6PD deficiency, demographic characteristics, and COVID-19 infection.

| Characteristic | Descriptive |  |  | p-value<sup>1</sup> | Univariate Regression |  |  | Multivariate Regression |  |  |
| --- | --- | --- | --- | --- | --- | --- | --- | --- | --- | --- |
|  | Total<br>N = 3,955 | No Covid<br>infection<br>N = 3,579 | Covid<br>infection<br>N = 376 |  | OR<sup>2</sup> | 95% CI<sup>2</sup> | p-value | OR<sup>2</sup> | 95% CI<sup>2</sup> | p-value |
| **G6PD deficient,**<br>**N (%)** | 206 (5.21%) | 185 (5.17%) | 21 (5.59%) | 0.7 | 1.09 | 0.66; 1.69 | 0.7 | 1.15 | 0.70; 1.79 | 0.6 |
| **Age, Average**<br>**(SD)** | 34.8 (16.9) | 34.3 (17.0) | 39.4 (15.3) | **<0.001** | 1.02 | 1.01; 1.02 | **<0.001** | 1.02 | 1.01; 1.02 | **<0.001** |
| **Race, n/N (%)** |  |  |  | **<0.001** |  |  |  |  |  |  |
| White | 259 (6.55%) | 236 (6.59%) | 23 (6.12%) |  | — | — |  | — | — |  |
| Black | 612 (15.47%) | 553 (15.45%) | 59 (15.69%) |  | 1.09 | 0.67; 1.85 | 0.7 | 1.01 | 0.61; 1.71 | >0.9 |
| Asian | 125 (3.16%) | 97 (2.71%) | 28 (7.45%) |  | 2.96 | 1.63; 5.44 | **<0.001** | 2.87 | 1.57; 5.29 | **<0.001** |
| Admixed | 2,899 (73.30%) | 2,643 (73.85%) | 256 (68.09%) |  | 0.99 | 0.65; 1.59 | >0.9 | 0.98 | 0.64; 1.57 | >0.9 |
| Indigenous | 60 (1.52%) | 50 (1.40%) | 10 (2.66%) |  | 2.05 | 0.89; 4.48 | 0.079 | 2.04 | 0.88; 4.46 | 0.083 |

<sup>1</sup> Pearson's Chi-squared test; Wilcoxon rank sum test
<sup>2</sup> OR = Odds Ratio, CI = Confidence Interval

No association was found between G6PD deficiency and hospitalization (OR = 1.13; 95% CI: 0.27–3.20; p = 0.8) or death (all 16 deaths were in the non-deficient group), even after adjustment for age and race. Age was strongly associated with both outcomes: for hospitalization, OR = 1.06 (p < 0.001); for death, OR = 1.06 (p < 0.001). No racial group was significantly associated with hospitalization or mortality in the adjusted models (Tables 3 and 4).

**Table 3.** Association between G6PD deficiency, demographic characteristics, and hospitalization due to COVID-19.

| Characteristic | Total<br>N = 3,955 | Descriptive |  | p-value<sup>1</sup> | Univariate Regression |  |  | Multivariate Regression |  |  |
| --- | --- | --- | --- | --- | --- | --- | --- | --- | --- | --- |
|  |  | Not hospitalized<br>N = 3,903 | Hospitalized<br>N = 52 |  | OR<sup>2</sup> | 95% CI<sup>2</sup> | p-value | OR<sup>2</sup> | 95% CI<sup>2</sup> | p-value |
| **G6PD deficient, N (%)** | 206 (5.21%) | 203 (5.20%) | 3 (5.77%) | 0.8 | 1.12 | 0.27; 3.07 | 0.9 | 1.13 | 0.27;3.20 | 0.8 |
| **Age, mean (SD)** | 34.8 (16.9) | 34.6 (16.8) | 51.6 (17.9) | **<0.001** | 1.06 | 1.04; 1.08 | **<0.001** | 1.06 | 1.04;1.08 | **<0.001** |
| **Race, N (%)** |  |  |  | 0.3 |  |  |  |  |  |  |
| White | 259 (6.55%) | 255 (6.53%) | 4 (7.69%) |  | — | — |  | — | — |  |
| Black | 612 (15.47%) | 604 (15.48%) | 8 (15.38%) |  | 0.84 | 0.26; 3.19 | 0.8 | 0.65 | 0.20;2.50 | 0.5 |
| Asian | 125 (3.16%) | 121 (3.10%) | 4 (7.69%) |  | 2.11 | 0.49; 9.05 | 0.3 | 1.97 | 0.45;8.65 | 0.4 |
| Admixed | 2,899 (73.30%) | 2,864 (73.38%) | 35 (67.31%) |  | 0.78 | 0.31; 2.62 | 0.6 | 0.78 | 0.30;2.64 | 0.6 |
| Indigenous | 60 (1.52%) | 59 (1.51%) | 1 (1.92%) |  | 1.08 | 0.05; 7.47 | >0.9 | 1.21 | 0.06;8.69 | 0.9 |

<sup>1</sup>Fisher's exact test; Wilcoxon rank sum test
<sup>2</sup>OR = Odds Ratio, CI = Confidence Interval

**Table 4.** Association between G6PD deficiency, demographic characteristics, and death from COVID-19.

| Characteristic | Descriptive |  |  |  | Univariate Regression |  |  | Multivariate Regression |  |  |
| --- | --- | --- | --- | --- | --- | --- | --- | --- | --- | --- |
|  | Total<br>N = 3,955 | No death<br>N = 3,939 | Death<br>N = 16 | p-value<sup>1</sup> | OR<sup>2</sup> | 95% CI<sup>2</sup> | p-value | OR<sup>2</sup> | 95% CI<sup>2</sup> | p-value |
| **G6PD deficient, N (%)** | 206 (5.21%) | 206 (5.23%) | 0 (0.00%) | >0.9 | 0.00 |  | >0.9 | 0.00 |  | >0.9 |
| **Age, mean (SD)** | 34.8 (16.9) | 34.8 (16.9) | 52.0 (17.4) | **<0.001** | 1.06 | 1.03; 1.09 | **<0.001** | 1.06 | 1.03; 1.09 | **<0.001** |
| **Race, N (%)** |  |  |  | 0.061 |  |  |  |  |  |  |
| White | 259 (6.55%) | 259 (6.58%) | 0 (0.00%) |  | — | — |  | — | — |  |
| Black | 612 (15.47%) | 606 (15.38%) | 6 (37.50%) |  | Inf<sup>4</sup> | 0.00; NA<sup>3</sup> | >0.9 | Inf<sup>4</sup> | 0.00; NA<sup>3</sup> | >0.9 |
| Asian | 125 (3.16%) | 125 (3.17%) | 0 (0.00%) |  | 1.00 | 0.00; Inf<sup>4</sup> | >0.9 | 0.85 | 0.00; Inf<sup>4</sup> | >0.9 |
| Admixed | 2,899 (73.30%) | 2,890 (73.37%) | 9 (56.25%) |  | Inf<sup>4</sup> | 0.00; NA<sup>3</sup> | >0.9 | Inf<sup>4</sup> | 0.00; NA<sup>3</sup> | >0.9 |
| Indigenous | 60 (1.52%) | 59 (1.50%) | 1 (6.25%) |  | Inf<sup>4</sup> | 0.00; NA<sup>3</sup> | >0.9 | Inf<sup>4</sup> | 0.00; NA<sup>3</sup> | >0.9 |

<sup>1</sup>Fisher's exact test; Wilcoxon rank sum test
<sup>2</sup>OR = Odds Ratio, CI = Confidence Interval
<sup>3</sup>NA= Not Applicable
<sup>4</sup>Inf= a very large numeric value

Kaplan–Meier curves (**Figure 3A–D**) were used to evaluate time to infection, hospitalization, ICU admission, and death by G6PD status. No significant differences were observed in any of the time-to-event outcomes, further supporting the absence of association between G6PD deficiency and adverse COVID-19 progression.

**Figure 3.**
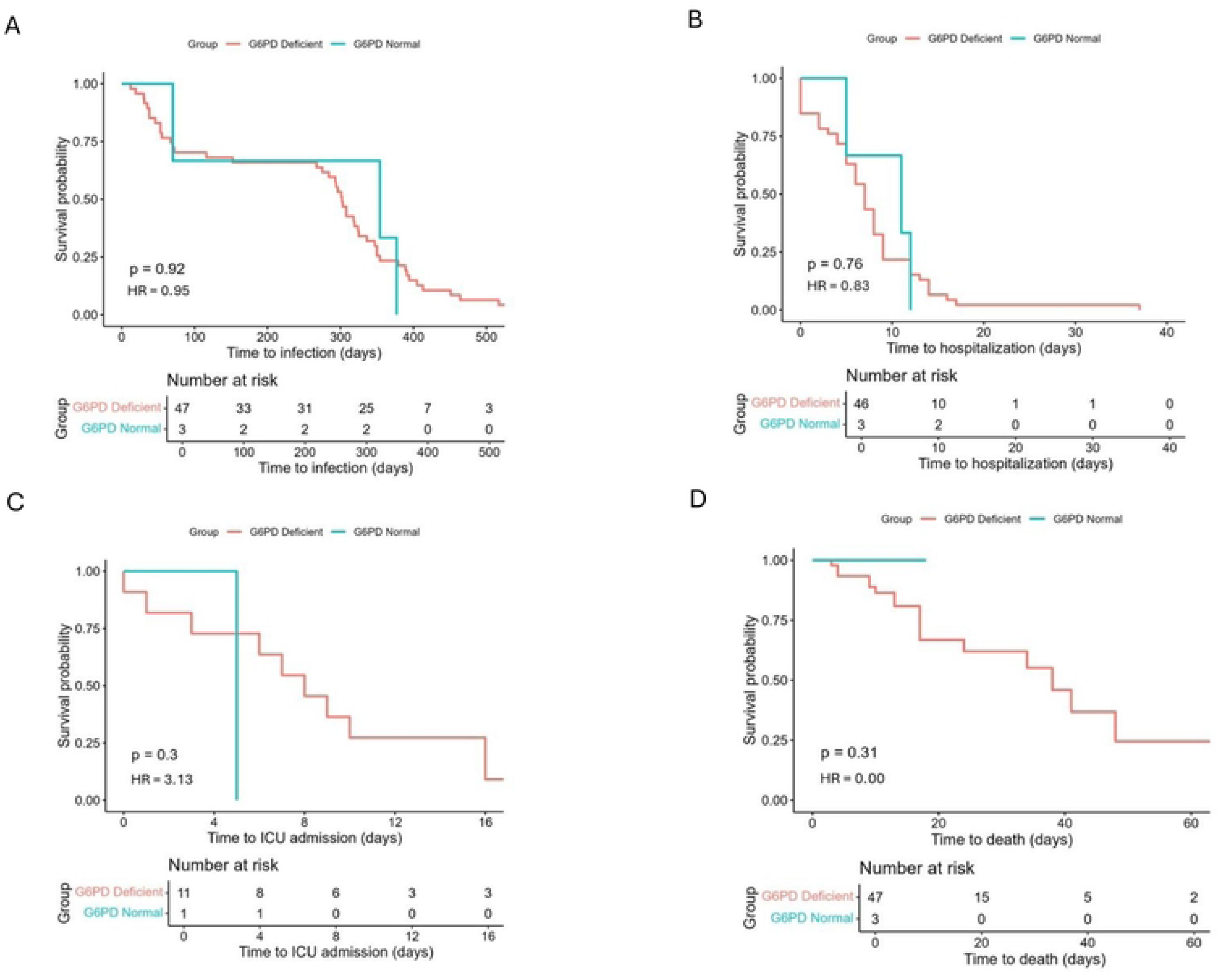
Kaplan–Meier curve comparing the probability of infection over time by G6PD status; A) Time to infection; B) Time to hospitalization; C) Time to ICU admission; D) Time to death in COVID-19 patients by G6PD status.

In addition to the main analysis, we performed two sensitivity analyses to assess the robustness of our findings. The first included only participants who had been screened for infectious diseases at the time of G6PD testing [31], aiming to minimize potential bias related to transient alterations in enzymatic activity. The second involved participants matched by G6PD status and age to account for potential confounding (**Supplementary Tables S1–S6**). In both subgroups, the results remained consistent with those of the full cohort, showing no significant associations between G6PD deficiency and COVID-19 outcomes. These findings support the internal validity of the study and suggest that the observed lack of association is unlikely to be due to sample composition or measurement bias.

## DISCUSSION

This retrospective cohort study evaluated the association between G6PDd and COVID-19 outcomes – infection, hospitalization, and mortality – in a male population whose enzymatic activity had been measured before the pandemic. Although early hypotheses suggested that G6PDd could worsen the clinical course of coronavirus infections due to its role in cellular oxidative stress responses [33,34], our findings did not demonstrate a statistically significant association between G6PDd and any of the evaluated outcomes in this population.

Specifically, G6PD-deficient (G6PDd) individuals were not at increased risk of SARS-CoV-2 infection, hospitalization, or death. These results are consistent with those of de Almeida et al. [23], who evaluated G6PD enzymatic activity and COVID-19 severity in a hospitalized cohort in the same region. That study also found no association between G6PDd and mortality or need for ventilatory support, although it did report a longer hospital stay for G6PDd patients. Together, these findings suggest that while G6PDd may influence some clinical aspects of care, it does not appear to directly impact the most severe outcomes of COVID-19.

A study conducted among United States veterans found an increased risk of severe COVID-19 in G6PD-deficient subgroups, particularly among Black veterans under 65 and White veterans aged 65 and older, suggesting that the relationship between G6PDd and disease severity may be modulated by age and ethnicity [20]. These data imply that G6PDd alone may not be a determinant of clinical severity but should be considered alongside other modifying factors in future analyses.

One potential explanation for the lack of association observed in our study relates to the predominant genetic variant of G6PD in the Brazilian Amazon, the G6PD*A− variant, and its repercussion on cell oxidative stress. This variant, classified as a Class III mutation, is associated with moderate enzymatic deficiency and partial preservation of redox capacity [31]. Such a mild phenotype may not be sufficient to impair antioxidant responses to the extent necessary to influence the clinical course of COVID-19. In contrast, Class I and II variants, which are more prevalent in the Mediterranean, Middle East, and parts of Asia, result in more severe enzymatic impairment and have been associated with increased vulnerability to infectious diseases. Studies suggest that it is the degree of enzymatic deficiency, rather than the mere presence of a G6PD mutation, that critically influences susceptibility to adverse outcomes, especially under conditions of heightened oxidative stress such as severe viral infections [35,36]. Additionally, the small number of severe events (hospitalizations and deaths) among G6PD-deficient individuals in our cohort may have limited the statistical power to detect meaningful associations. This further underscores the need for larger multicenter studies capable of stratifying participants by variant class and clinical severity.

The survival analyses of time-to-event outcomes (hospitalization, ICU admission, and death) also revealed no significant differences between G6PD-deficient and non-deficient groups. This temporal similarity supports the regression results and suggests that G6PDd does not impact disease progression. In contrast, age consistently emerged as a strong risk factor across all outcomes measured. This aligns with a broad body of literature demonstrating that advanced age is associated with worse COVID-19 outcomes regardless of other risk factors [37–40].

Another relevant finding was the significant association between Asian ethnicity and an increased risk of SARS-CoV-2 infection. This observation is supported by studies such as that by Sze et al. [41], which found heightened COVID-19 susceptibility among individuals of Asian descent in the UK, even after adjusting for socioeconomic factors. While racial analyses in Brazil traditionally focus on disparities affecting Black and Admixed populations [42], our findings underscore the importance of expanding vulnerability assessments to include other ethnic groups, particularly in demographically diverse regions such as the Amazon.

The increased risk observed among Asian-descendant participants in our study mirrors available evidence, where higher infection and hospitalization rates were noted among Asian populations [43]. These disparities are likely multifactorial, potentially reflecting occupational exposure, population density, genetic predisposition, or differential access to healthcare. However, in the Brazilian context, such associations remain underexplored. Further research is warranted to better understand the determinants of risk in Asian-descendant groups and ensure their adequate representation in public health surveillance and policy development.

The lack of association between G6PDd and severe COVID-19 may also reflect phenotypic variability [17]. Clinical expression of G6PDd likely depends on interactions with contextual oxidative stressors such as certain drugs (e.g., sulfonamides and primaquine) or coinfections, which may not have been present or prevalent in our cohort [44].

Our sensitivity analyses support the robustness of the findings. These analyses included participants who tested negative for malaria and were afebrile at the time of G6PD testing, aiming to avoid enzymatic activity modulation by acute infections [45,46]. Because phenotypic tests measure enzymatic activity rather than genotype, the exclusion of participants with active infections was intended to reduce potential misclassification. Additionally, we performed age-matched subgroup analyses to mitigate sample distribution bias, given the 5.2% G6PDd prevalence. The consistency of these results with the main findings further reinforces the internal validity of the study.

This study has some limitations, including the lack of data on viral load, comorbidities, and vaccination status; the potential underreporting of mild or asymptomatic cases; the restriction to male participants, due to the methodology used to identify complete G6PD deficiency; and the fact that G6PD testing was not performed during the pandemic. One notable limitation is the absence of granular clinical data, such as the need for mechanical ventilation or laboratory markers of severity, which have been used in other studies to capture intermediate clinical outcomes. Although we evaluated hospitalization and mortality, we were unable to assess whether G6PD deficiency influenced other severe outcomes. These indicators could have provided additional insights into the potential role of G6PD in modulating the severity of the inflammatory response or hypoxia-related injury in COVID-19 in the local context. Also, although G6PD enzymatic activity was assessed pre-pandemically, the absence of molecular genotyping limits the ability to evaluate variant-specific effects, particularly for rare Class I and II mutations that may confer higher clinical risk. Finally, the relatively small number of hospitalizations and deaths among G6PD-deficient individuals reduced the statistical power to detect associations with severe COVID-19 outcomes.

## CONCLUSION

Our findings indicate that G6PD deficiency was not associated with increased risk of infection, hospitalization, UCI admission, or death from COVID-19. Age remained the strongest predictor of adverse outcomes.

## Data Availability

All relevant data are within the manuscript and its Supporting Information files

## ACKNOWLEDGMENTS

We acknowledge the Brazilian Ministry of Health, through the Unified Health System (SUS), and the professionals responsible for maintaining the health databases that made this study possible. We are also grateful to the patients whose data contributed to these findings.

## SUPPORTING INFORMATION

**S1. Table:** Descriptive and regression sensitivity analysis of COVID-19 incidence in individuals with and without G6PD deficiency. Only participants who had been screened for infectious diseases at the time of G6PD testing were included. (DOCX)

**S2. Table:** Descriptive and regression sensitivity analysis of COVID-19 hospitalization in individuals with and without G6PD deficiency. Only participants who had been screened for infectious diseases at the time of G6PD testing were included. (DOCX)

**S3. Table:** Descriptive and regression sensitivity analysis of death from COVID-19 in individuals with and without G6PD deficiency. Only participants who had been screened for infectious diseases at the time of G6PD testing were included. (DOCX)

**S4. Table:** Descriptive and regression sensitivity analysis of COVID-19 infection in a subsample matched by age. (DOCX)

**S5. Table:** Descriptive and regression sensitivity analysis of hospitalization due to COVID-19 in a sample matched by age. (DOCX)

**S6. Table:** Descriptive and regression sensitivity analysis of death from COVID-19 in a sample matched by age. (DOCX)

